# Novel Machine-Learned Approach for COVID-19 Resource Allocation: A Tool for Evaluating Community Susceptibility

**DOI:** 10.1101/2020.10.14.20212571

**Authors:** Neil Kale

## Abstract

Despite worldwide efforts to develop an effective COVID vaccine, it is quite evident that initial supplies will be limited. Therefore, it is important to develop methods that will ensure that the COVID vaccine is allocated to the people who are at major risk until there is a sufficient global supply. Herein, I developed a machine-learning tool that could be applied to assess the risk in communities based on social, medical, and lifestyle risk factors. As a “proof of concept,” I modeled COVID risk in the Massachusetts communities using 29 risk factors, including the prevalence of preexisting comorbid conditions like COPD and social factors such as racial composition. Of the 29 factors, 14 were found to be significant (p < 0.1) indicators: poverty, food insecurity, lack of high school education, lack of health insurance coverage, premature mortality, population, population density, recent population growth, Asian percentage, high-occupancy housing, and preexisting prevalence of cancer, COPD, overweightness, and heart attacks. The machine-learning approach finds the 9 highest risk communities in the state of Massachusetts: Lynn, Brockton, Revere, Randolph, Lowell, New Bedford, Everett, Waltham, and Fitchburg. The 5 most at-risk counties are Suffolk, Middlesex, Bristol, Norfolk, and Plymouth. With appropriate data, the tool could evaluate risk in other communities, or even enumerate individual patient susceptibility. A ranking of communities by risk may help policymakers ensure equitable allocation of limited doses of the COVID vaccine.

## Introduction

The COVID-19 pandemic has had an uneven impact worldwide. Just as certain patients are at higher risk of severe illness, such as the elderly and those with preexisting conditions, entire communities can exhibit a disproportionately high susceptibility to infection. Indeed, dozens of community-wide risk factors have been found, ranging from demographics to environmental factors such as air pollution (Wu et al., 2020). Clinical risk scores for patients are in development since spring (Liang et al., 2020). They are used in hospitals around the country to allocate ventilators. More recently, scores for certain community sets such as American counties (Peters, 2020) or large American cities (Gourevitch et al., 2020) have emerged. Because initially COVID vaccine supply will be limited, it is important to ensure that vaccines are allocated to the communities that are at high risk. Herein, I developed a machine-learning tool that could be applied to Massachusetts communities and can be applied to assess the risk in other communities based on social, medical, and lifestyle risk factors.

## Methodology

### Data Collection

Data by town for the 29 potential risk factors was found online in the public domain. Refer to Table 1 for a list of sources, and Appendix A for the complete dataset. To minimize covariance, incidence of preexisting conditions, number of COVID cases, number of residents by race, number of residents by age, number of residents on SNAP, and town revenue were adjusted for population. The explanatory variables were grouped into three categories: medical, demographic, and economic (See row 2, Appendix A). The population density was calculated by iterating through the MA Land Parcel Database to sum up the land area of each town, then dividing the population by land area. The moving rate was calculated as (100 - % of residents who lived in the same house in the last year).

### Model

Binary logistic regression was used to fit models to the data. This method is highly flexible, allowing the tool to handle categorical or continuous independent variables. Furthermore, unlike KNN or decision tree models, logistic regression returns an easily-interpretable numeric result. Towns were assigned ‘high’ or ‘low’ with respect to the median COVID cases per 1,000 residents. This ensured balanced classes. The model returned a value usually within 0 to 1, where one or above means confident that a town is at high risk, and zero or below means confident that a town is at low risk. This value was interpreted as the town’s risk score. A separate model was trained on each category of predictors, and on all the predictors at once. Finally, the optimal model was trained on the significant predictors (p < 0.10) from all the previous models. The initial models only contained an intercept and linear term for each predictor. All predictors in the optimal model were verified as correlated to COVID risk, so the optimal model added terms for the square of each predictor (MathWorks). Instructions on how to install an interactive version of the model are provided at https://blog.sunnysideup.life/?p=198.

## Results

The optimal model with quadratic terms is 81.5% accurate on the training data. The other models are less accurate, see Appendix B for values. MATLAB Classification Learner found the optimal model (only linear terms because of software limitations) as 75.2% accurate with 5-fold cross-validation. As shown in Figure 1, the misclassification rate for the optimal model (linear terms only) is approximately equal for the high and low classes, and as in Figure 2, the AUC is 0.81. Suffolk County has the highest average risk score and the highest percentage of high-risk towns, see Table 2. The same data is displayed graphically in Figures 3-5. Fourteen of the COVID risk factors were found to be significant (p < 0.10), see Table 3. In the optimal model, where factors were selected from pairs of, squares of, and individual factors that were found to be significant in earlier models, 9 factors were found to be significant, see Table 4.

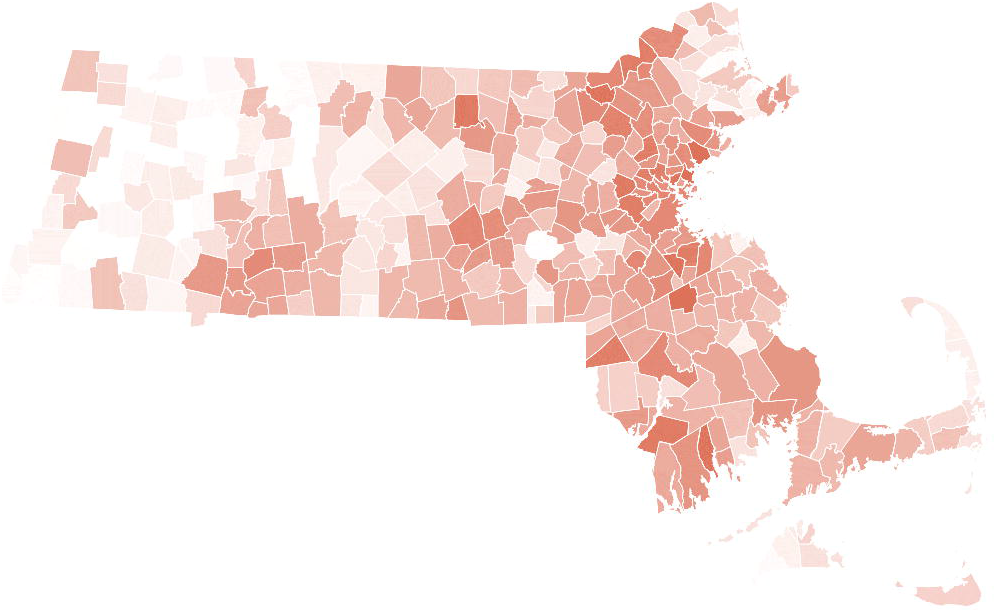

## Discussion

The novel machine-learning model developed herein found population density to be a significant factor (p = 0.026) in determining COVID risk in MA towns. In contrast, Johns Hopkins researchers found it not significant (Hamidi et al., 2020) suggesting that further research is required to determine the effect of population density on COVID risk. The 7 potential medical factors were all chosen based on prior research (Zheng et al., 2020); however, the model suggests that, out of those, heart attacks per 10,000 people (p = 0.014) is the most significant COVID risk factor for communities. While COPD rate and overweight percentage were significant in the medical model, only heart attack rate was significant in the optimal model. While 29 factors were collected for this study, community risk scoring might be accomplished with many subsets of that data. For example, the medical model, which used only preexisting conditions and mortality rates, achieved 75.5% accuracy on the training data. The demographic model used a completely disjoint subset of the risk factors and achieved 77.2% accuracy on the training data. The significance of Asian percentage as an indicator of community COVID risk is somewhat surprising and merits further research. Two possible explanations are that, for medical reasons, Asians are more prone to infection by COVID, or that social considerations may increase COVID susceptibility in communities with a high Asian population.

## Conclusion

In summary, the results of this model will be useful in guiding further research into community susceptibility to COVID. Such research is of paramount importance. As COVID vaccines become available beyond healthcare providers and essential workers, they can be allocated to the highest-risk communities, thus preventing the maximum number of future COVID cases. The approach used herein can be extrapolated for analysis of other states and countries, so that governing bodies at those respective levels can be aided in forming a localized vaccination plan.

## Supporting information

Appendix B: Supplemental Results

## Data Availability

The author confirms that the data supporting the findings of this study are available within the article and its supplementary materials.

