## Supplementary material for "Novel Machine-Learned Approach for COVID-19 Resource Allocation: A Tool for Evaluating Community Susceptibility": Tables and Figures

Table 1. Data sources.

| **Risk Factor** | **Link to Source** |
|---|---|
| COVID-19 Cases (Municipal) | https://www.mass.gov/info-details/covid-19-response-reporting#covid-19-weekly-public-health-report- |
| Population Estimates 2010 - 2019 (Municipal) | http://www.donahue.umassp.edu/business-groups/economic-public-policy-research/massachusetts-population-estimates-program/population-estimates-by-massachusetts-geography/by-city-and-town |
| Foodstamps/SNAP Households by Kids & HH Type (Municipal) | https://datacommon.mapc.org/browser/datasets/143 |
| Population in Poverty (Municipal) | https://datacommon.mapc.org/browser/datasets/57 |
| Population with Health Insurance by Gender and Age (Municipal) | https://datacommon.mapc.org/browser/datasets/204 |
| COPD, Heart Attack Cases (Municipal) | https://matracking.ehs.state.ma.us/ |
| Cancer, Diabetes, Obesity, Overweight Cases (Municipal) | https://www.mass.gov/orgs/population-health-information-tool-phit |

Table 2. Average COVID-19 risk and % high-risk towns by Massachusetts county.

| **County** | **Total Risk** | **Num. Towns** | **Avg. Risk** | **Num. High Risk Towns** | **% High Risk Towns** |
|---|---|---|---|---|---|
| Suffolk | 3.81445 | 4 | **0.95** | 4 | **100%** |
| Middlesex | 40.04917 | 54 | **0.74** | 43 | **80%** |
| Bristol | 14.81407 | 20 | **0.74** | 16 | **80%** |
| Norfolk | 20.12683 | 28 | **0.72** | 24 | **86%** |
| Plymouth | 16.89145 | 27 | **0.63** | 22 | **81%** |
| Essex | 19.34298 | 34 | **0.57** | 19 | **56%** |
| Worcester | 30.56389 | 60 | **0.51** | 32 | **53%** |
| Hampden | 11.35392 | 23 | **0.49** | 12 | **52%** |
| Barnstable | 6.24452 | 15 | **0.42** | 6 | **40%** |
| Nantucket | 0.39544 | 1 | **0.40** | 0 | **0%** |
| Hampshire | 5.601604 | 20 | **0.28** | 6 | **30%** |
| Dukes | 1.170723 | 7 | **0.17** | 0 | **0%** |
| Franklin | 2.566782 | 26 | **0.10** | 2 | **8%** |
| Berkshire | 3.064332 | 32 | **0.10** | 3 | **9%** |

Table 3. Significant risk factors, with p-value, and the model where each was found significant. "All" signifies the model with all risk factors, and economic, demographic, and medical signify the model with that subgrouping of factors. Strength is calculated as the estimated coefficient multiplied by the average value, and it compares how each of the risk factors affects the risk score.

| **Risk Factor** | **p-Value** | **Model** | **Strength** |
|---|---|---|---|
| Poverty Rate | 0.005 | All | -0.214 |
| % Without Health Insurance Coverage | 0.065 | All | -0.086 |
| Premature Mortality per 100,000 | 0.066 | All | 0.181 |
| Population | 0.003/0.000 | All/Demographic | 0.135 / 0.227 |
| Asian % | 0.061/0.047 | All/Demographic | 0.069 / 0.070 |
| Num. Housing Facilities with Over 20 Residents | 0.002/0.000 | All/Demographic | -0.058 / -0.100 |
| Cancer Case per 100,000 | 0.041 | All | -0.104 |
| COPD per 100,000 | 0.011/0.017 | All/Medical | 0.080 / 0.071 |
| Heart Attacks per 100,000 | 0.025/0.001 | All/Medical | 0.082 / 0.119 |
| Overweight % | 0.063/0.021 | All/Medical | 0.351 / 0.406 |
| Population Density | 0.076 | Demographic | 0.035 |
| % Less than High School Education | 0.053 | Demographic | 0.096 |
| % Change in Population | 0.001 | Demographic | 0.083 |
| Household Snap Rate | 0.000 | Economic | 0.244 |

Table 4. Significant risk factors in the optimal model, along with p-values and strengths for each factor.

| **Risk Factor** | **p-Value** | **Strength** |
|---|---|---|
| AsianPcnt | 0.013 | 0.135 |
| HeartAttackPer10_000 | 0.014 | 0.226 |
| PcntChangeInPop | 0.012 | 0.091 |
| Pop | 0.026 | 0.188 |
| PopulationDensity | 0.056 | 0.086 |
| AsianPcnt^2 | 0.044 | -0.016 |
| HeartAttackPer10_000^2 | 0.095 | -0.055 |
| PcntChangeInPop^2 | 0.004 | -0.028 |
| Pop^2 | 0.089 | -0.014 |

Figures

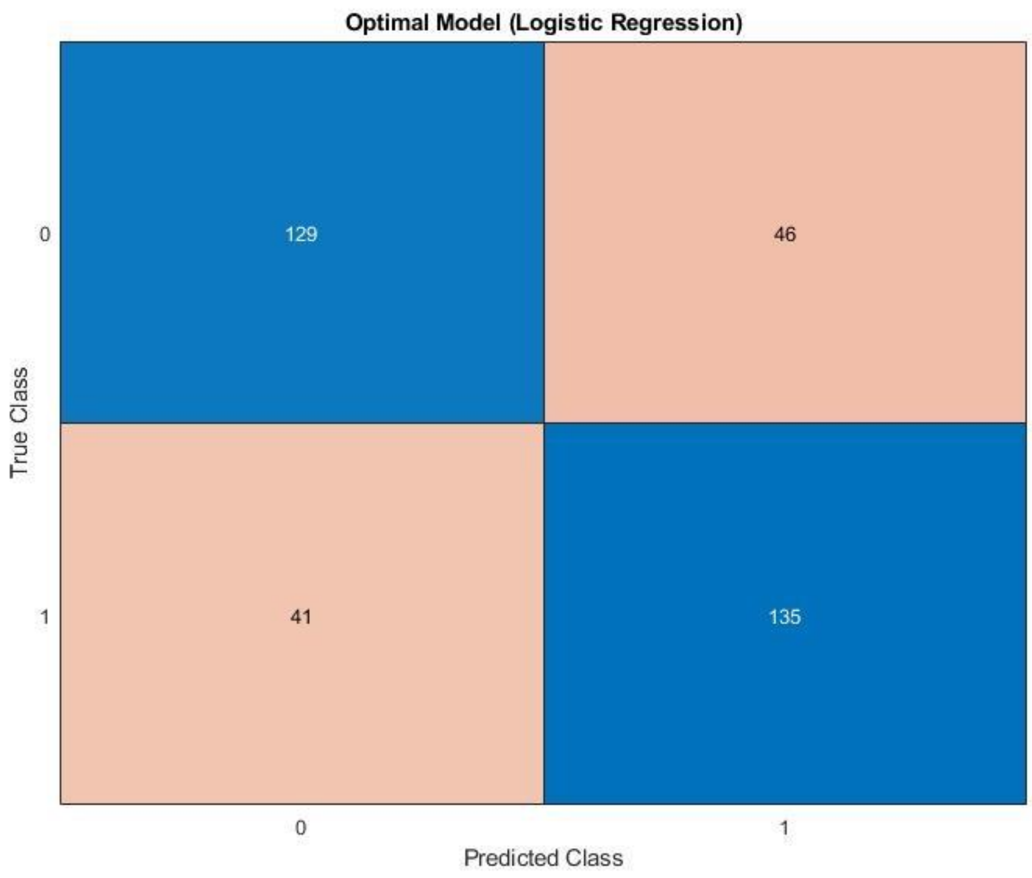


Figure 1. Optimal model (linear terms only) confusion matrix.

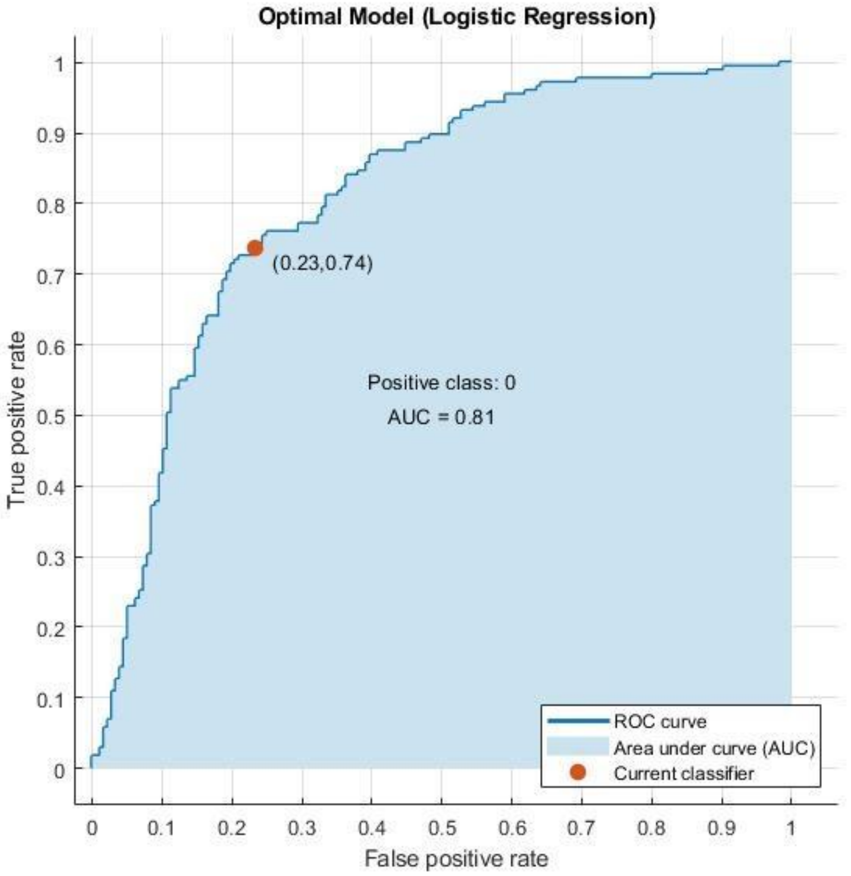


Figure 2. Optimal model (linear terms only) ROC curve.

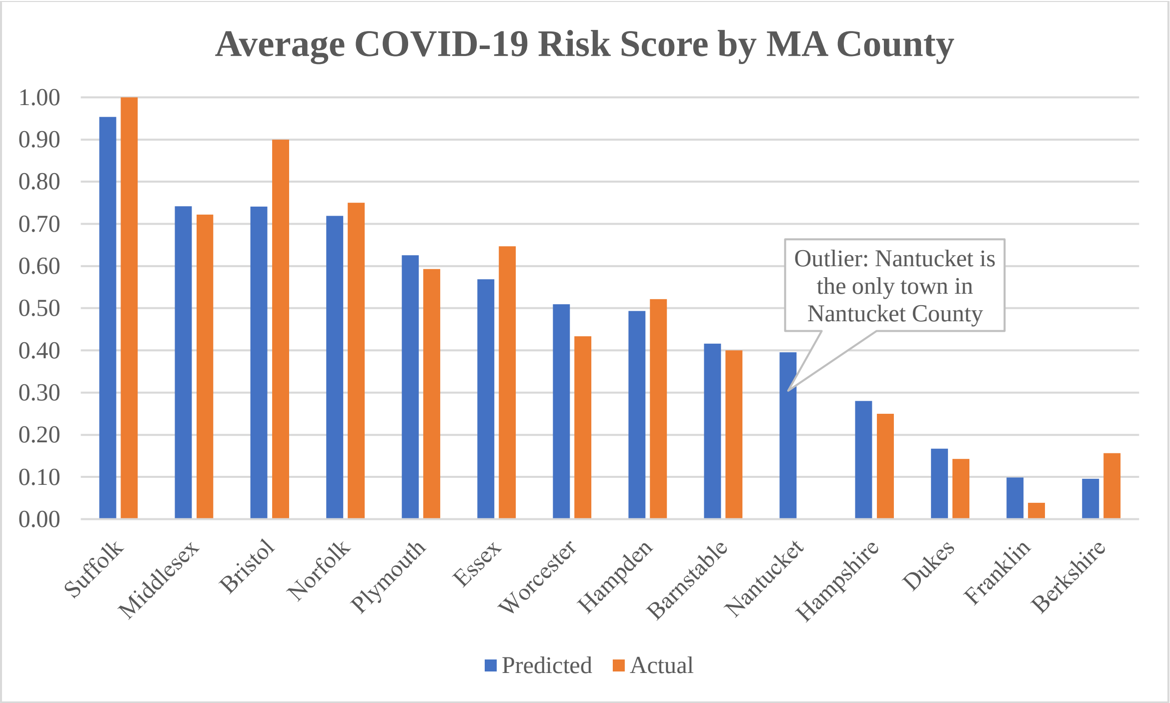


Figure 3. Average COVID-19 risk score by MA county.

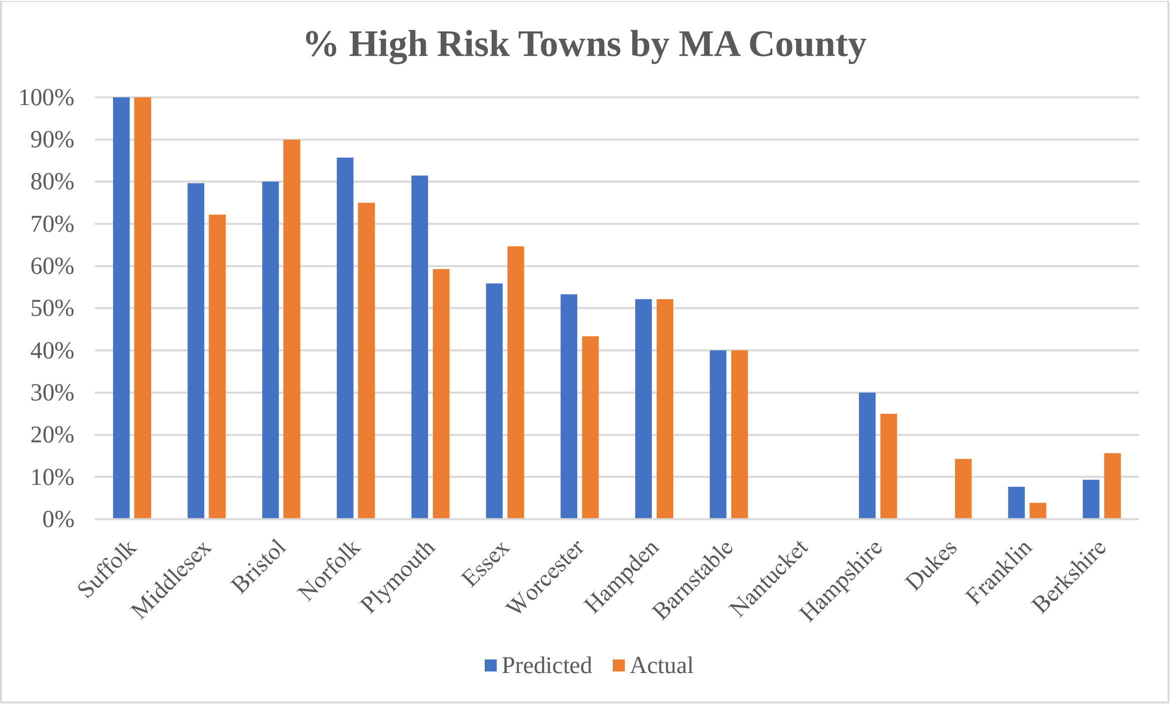


Figure 4. Percentage of high risk towns by MA county.

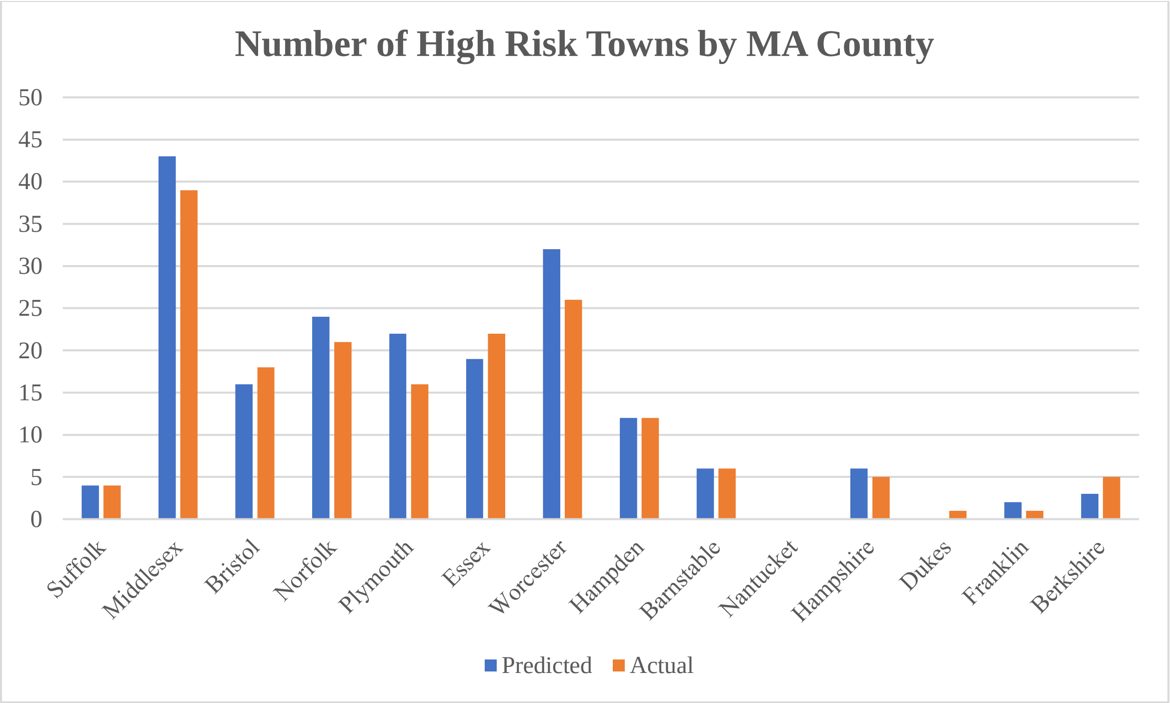


Figure 5. Total number of high risk communities by MA county.
